# Cardiovascular Disease Risk Prediction Models in Haiti: Implications for Primary Prevention in Low-Middle Income Countries

**DOI:** 10.1101/2021.07.29.21261344

**Authors:** Lily D Yan, Jean Lookens Pierre, Vanessa Rouzier, Michel Théard, Alexandra Apollon, Stephano St-Preux, Justin R Kingery, Kenneth A Jamerson, Marie Deschamps, Jean W Pape, Monika M Safford, Margaret L McNairy

**Affiliations:** Division of General Internal Medicine, Department of Medicine, Weill Cornell Medicine, New York, New York, USA; Center for Global Health, Weill Cornell Medicine, New York, New York, USA; Haitian Group for the Study of Kaposi’s Sarcoma and Opportunistic Infections (GHESKIO), Port-au-Prince, Haiti; Collège Haïtien de Cardiologie, Port-au-Prince, Haiti; Division of Cardiovascular Disease, Department of Medicine, University of Michigan, Ann Arbor, Michigan, USA

**Keywords:** cardiovascular diseases, primary prevention, risk, global health, hypertension

## Abstract

**Background:** Cardiovascular diseases (CVD) are rapidly increasing in low-middle income countries (LMICs). Accurate risk assessment is essential to reduce premature CVD by targeting primary prevention and risk factor treatment among high-risk groups. Available CVD risk prediction models are built on predominantly Caucasian, high-income country populations, and have not been evaluated in LMIC populations.

**Objective:** To compare the predicted 10-year risk of CVD and identify high-risk groups for targeted prevention and treatment in Haiti.

**Methods:** We used cross-sectional data within the Haiti CVD Cohort Study, including 653 adults ≥ 40 years without known history of CVD and with complete data. Six CVD risk prediction models were compared: pooled cohort equations (PCE), adjusted PCE with updated cohorts, Framingham CVD Lipids, Framingham CVD Body Mass Index (BMI), WHO Lipids, and WHO BMI. Risk factors were measured during clinical exams. Primary outcome was continuous and categorical predicted 10-year CVD risk. Secondary outcome was statin eligibility.

**Results:** Seventy percent were female, 65.5% lived on a daily income of ≤1 USD, 57.0% had hypertension, 14.5% had hypercholesterolemia, 9.3% had diabetes mellitus, 5.5% were current smokers, and 2.0% had HIV. Predicted 10-year CVD risk ranged from 3.9% in adjusted PCE (IQR 1.7-8.4) to 9.8% in Framingham-BMI (IQR 5.0-17.8), and Spearman rank correlation coefficients ranged from 0.87 to 0.98. The percent of the cohort categorized as high risk using the uniform threshold of 10-year CVD risk ≥ 7.5% ranged from 28.8% in the adjusted PCE model to 62.0% in the Framingham-BMI model (χ^2^ = 331, p value < 0.001). Statin eligibility also varied widely.

**Conclusions:** In the Haiti CVD Cohort, there was substantial variation in the proportion identified as high-risk and statin eligible using existing models, leading to very different treatment recommendations and public health implications depending on which prediction model is chosen. There is a need to design and validate CVD risk prediction tools for low-middle income countries that include locally relevant risk factors.

**Trial registration:** clinicaltrials.gov NCT03892265

## Background

Cardiovascular diseases (CVD) are rapidly increasing in low-middle income countries (LMICs), with ischemic heart disease, stroke, and peripheral artery disease amounting to over 17 million deaths in 2017 (1). Furthermore, mortality and disability due to CVD have increased by 21.1% and 16.4%, respectively, over the past ten years (1). Multiple risk factors contribute to the increase in CVD, including high systolic blood pressure (SBP), hyperlipidemia, tobacco and alcohol use (2). In addition to known modifiable risk factors such as diet, lack of physical activity, and smoking, many LMICs may have additional structural factors for CVD risk such as heavy metal environmental pollution and increased allostatic load related to the stress of poverty that are not captured in existing CVD risk prediction models (3,4).

Accurate CVD risk prediction to target use of statins and antihypertensives in primary CVD prevention is essential to reduce premature disease, especially in a LMIC country like Haiti where CVD leads mortality at 26.5% of all adults deaths, and where there are significant resource-constraints. (5,6). However, available CVD risk prediction models are built on predominantly Caucasian, high-income country populations, and have not been evaluated in LMIC populations due to the paucity of rigorous cohorts with hard CVD outcomes (7–9). Furthermore, model choice may have ramifications for which individuals are identified as high risk and recommended for treatment, with divergent cost and public health implications.

The aim of this study is to compare the estimated 10-year risk of CVD across six commonly used CVD prediction models, and identify high-risk groups for targeted statins. By applying these models to a population-based cohort in Haiti, we hope to inform which CVD risk prediction model might best be used in a LMIC context.

## Methods

### Study Design

We used cross-sectional data within the Haiti CVD Cohort Study, a population-based cohort in Port-au-Prince selected using multistage random sampling with a previously described protocol (clinicaltrials.gov NCT03892265) (10). We included 776 adults ≥ 40 years enrolled between March 2019 to April 2020. Participants already on a statin (n = 11), with history of angina, myocardial infarction, stroke (n= 69), or with missing data required for risk prediction (n = 72) were excluded given the focus on primary prevention (Supplemental Figure 1).

The study was conducted at the Groupe Haïtien d’Etude du Sarcome de Kaposi et des Infections Opportunistes clinics (GHESKIO), a medical organization that has operated continuously over four decades in Haiti to provide clinical care and conduct research on HIV and chronic diseases.

### Measurements

Demographic data (age, sex, education, income) and health behaviors (smoking status, physical activity) were collected during an enrollment survey using standardized WHO STEPs instruments (11). Clinical data, including height, weight, and blood pressure (BP), were measured during a physical exam with a study physician or nurse at enrollment.

BP was measured using the automated Omron HEM-907 machine with an appropriate cuff size (bladder encircling at least 80% of arm), after the participant had been seated in a quiet space for five minutes with both feet on the ground and their arm supported at heart level (11,12). Three BP measurements were taken on the left arm separated by one-minute intervals. In accordance with WHO guidelines, the second and third BP measurements were averaged for all analyses (11).

Medical history and diagnoses (hypertension, hyperlipidemia, diabetes, myocardial infarction, angina, stroke, HIV) were determined based a history and physical exam performed by a trained study physician, and direct clinical or laboratory measurement where applicable (Supplemental Table 1).

### CVD Risk Assessment

Six models were compared: the Pooled Cohort Equations (PCEs) (7), an adjusted PCE (aPCE) incorporating updated cohorts with more African Americans (13), Framingham CVD Lipids (8), Framingham CVD Body Mass Index (BMI) (8), WHO-Lipids (9), and WHO-BMI (9). These models were chosen because they are widely used, frequently compared in existing literature, and most include people of African descent. The systematic coronary risk evaluation (SCORE) model based on European cohorts was not used given it only predicts fatal CVD outcomes.

Underlying equations and coefficients were extracted from published literature and applied to the cohort (7–9,13) (Supplemental Table 2-4).

### Outcomes and Statistical Analysis

The primary outcome was predicted 10-year risk of CVD as 1) a continuous score and 2) a categorical score (low, intermediate, high). Categorical scores were calculated using two methods: uniform thresholds (low <5%, intermediate 5 to 7.5%, high ≥ 7.5%), and model specific thresholds (PCE and adjusted PCE: <5%, 5 to 7.5%, ≥ 7.5%; Framingham-Lipids and Framingham-BMI: <10%, 10 to 20%, ≥ 20%; WHO-Lipids and WHO-BMI: <5%, 5 to 20%, ≥ 20%). Model specific thresholds exist due to differences in equation derivation, including measured CVD outcomes (7–9). Spearman rank correlation coefficients were used to measure concordance between models’ ranked order of participants from lowest to highest risk, ranging from -1 (perfect discordance) to +1 (perfect concordance). Categorical scores were compared using chi square tests of independence. Discordance was defined as participants categorized as low risk by one score, but high risk by another.

For participants categorized as high-risk using uniform thresholds, the underlying risk factors were summarized using medians, counts, and percentages to understand what risk factors were leading to the high-risk scores.

The secondary outcome was statin eligibility, based on model specific thresholds and criteria (Supplemental Table 5). Statin eligibility was compared using chi square tests of independence. 95% confidence intervals (CI) were calculated using one sample proportions test.

All analyses were conducted using R, version 4.0.2.

### Results

Out of 1,435 adults ≥ 18 years enrolled during the study period, 1363 (95.0%) had complete data and 653 (45.5%) met study eligibility criteria. Of these 653, 70.4% were female, 65.5% lived on a daily income of ≤1 USD, 57.0% had hypertension, 14.5% had hypercholesterolemia, 9.3% had diabetes mellitus, 5.5% were current smokers, and 2.0% had HIV (Table 1). Overall, 38.1% had a low-density lipoprotein cholesterol greater than 130 mg/dL, 42.7% had a systolic blood pressure ≥ 140 mmHg, and 26.6% had a diastolic blood pressure ≥ 90 mmHg.

**Table 1:** Demographic and clinical characteristics of Haiti CVD Cohort (N = 653).

|  | N (%), or median [IQR] |
| --- | --- |
| **Female** | 460 (70.4) |
| **Age**, median [IQR], y | 54 [47, 62] |
| **Education**, primary or lower | 391 (59.9) |
| **Works for pay** | 231 (35.4) |
| **Income (daily)**, ≤1 USD | 428 (65.5) |
| **Comorbidities\*** |  |
| Hypertension | 372 (57.0) |
| On treatment | 143 (21.9) |
| Hypercholesterolemia | 95 (14.5) |
| On treatment | 0 |
| Diabetes Mellitus | 61 (9.3) |
| On treatment | 32 (4.9) |
| HIV | 13 (2.0) |
| **Smoking**, current | 36 (5.5) |
| **Physical Activity**, ≤ 150 min / week (low) | 370 (56.7) |
| **Alcohol intake**, more than 1 drink a day (moderate-high) | 9 (1.4) |
| **BMI ≥ 30 kg/m<sup>2</sup>** | 155 (23.7) |
| **Cholesterol** |  |
| HDL Cholesterol < 40 mg/dL | 137 (21.0) |
| LDL Cholesterol ≥130 mg/dL | 249 (38.1) |
| **Blood Pressure** |  |
| SBP ≥ 140 mmHg | 279 (42.7) |
| SBP ≥ 130 mmHg | 373 (57.1) |
| DBP ≥ 90 mmHg | 174 (26.6) |
| DBP ≥ 80 mmHg | 326 (49.9) |

Legend: IQR = interquartile range. \* Definitions available in Supplement.

### Predicted 10-year CVD Risk

Using a continuous score, median predicted 10-year CVD risk ranged from 3.9% in the adjusted PCE model (IQR 1.7-8.4) to 9.8% in the Framingham-BMI model (IQR 5.0-17.8) (Table 2). Using the Spearman rank correlation coefficient, we assessed the concordance between how each model ranked each individual participant in order from lowest risk to highest risk in pairwise comparisons. Spearman coefficients showed high concordance between models, ranging from 0.87 (Framingham-Lipids vs WHO-BMI) to 0.98 (PCE vs adjusted PCE).

However, categorization of individuals into risk groups using uniform thresholds showed extremely wide variability. The percent of the cohort categorized as high-risk ranged from 28.8% in the adjusted PCE model to 62.0% in the Framingham-BMI model (χ^2^ = 331, p value < 0.001) (Fig 1A). Under uniform thresholds, 186 participants had discordant scores (categorized as high risk by one score, but low risk by another). The most common pattern was categorization as high risk by Framingham-lipids or Framingham-BMI and low risk by another model (185 out of 186 discordant participants).

**Table 2:**
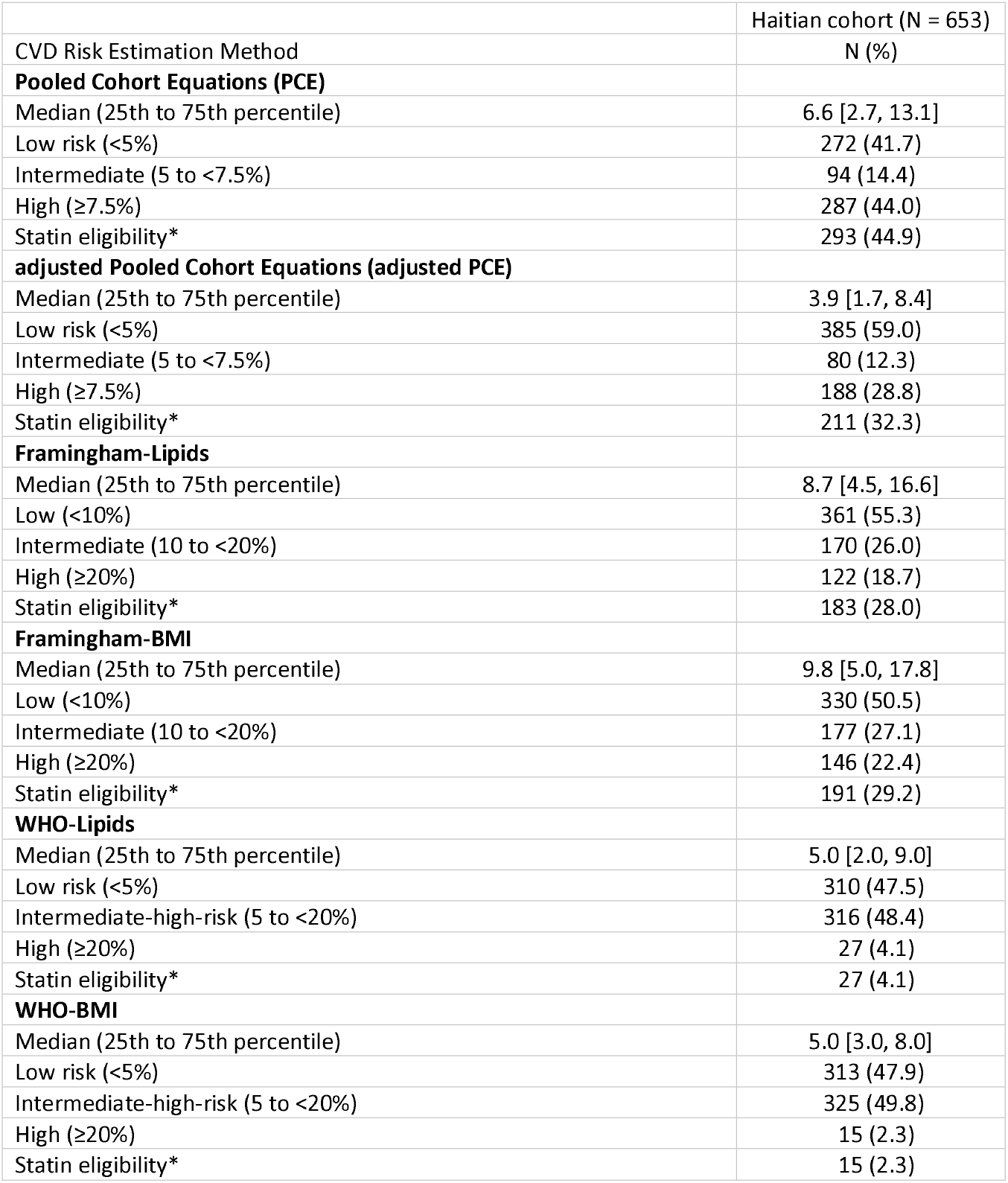
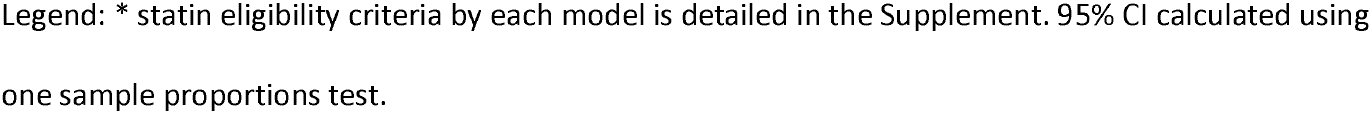
Predicted 10-year CVD risk in Haiti CVD Cohort.

| CVD Risk Estimation Method | Haitian cohort (N = 653)<br>N (%) |
| --- | --- |
| **Pooled Cohort Equations (PCE)** |  |
| Median (25th to 75th percentile) | 6.6 [2.7, 13.1] |
| Low risk (<5%) | 272 (41.7) |
| Intermediate (5 to <7.5%) | 94 (14.4) |
| High (≥7.5%) | 287 (44.0) |
| Statin eligibility* | 293 (44.9) |
| **adjusted Pooled Cohort Equations (adjusted PCE)** |  |
| Median (25th to 75th percentile) | 3.9 [1.7, 8.4] |
| Low risk (<5%) | 385 (59.0) |
| Intermediate (5 to <7.5%) | 80 (12.3) |
| High (≥7.5%) | 188 (28.8) |
| Statin eligibility* | 211 (32.3) |
| **Framingham-Lipids** |  |
| Median (25th to 75th percentile) | 8.7 [4.5, 16.6] |
| Low (<10%) | 361 (55.3) |
| Intermediate (10 to <20%) | 170 (26.0) |
| High (≥20%) | 122 (18.7) |
| Statin eligibility* | 183 (28.0) |
| **Framingham-BMI** |  |
| Median (25th to 75th percentile) | 9.8 [5.0, 17.8] |
| Low (<10%) | 330 (50.5) |
| Intermediate (10 to <20%) | 177 (27.1) |
| High (≥20%) | 146 (22.4) |
| Statin eligibility* | 191 (29.2) |
| **WHO-Lipids** |  |
| Median (25th to 75th percentile) | 5.0 [2.0, 9.0] |
| Low risk (<5%) | 310 (47.5) |
| Intermediate-high-risk (5 to <20%) | 316 (48.4) |
| High (≥20%) | 27 (4.1) |
| Statin eligibility* | 27 (4.1) |
| **WHO-BMI** |  |
| Median (25th to 75th percentile) | 5.0 [3.0, 8.0] |
| Low risk (<5%) | 313 (47.9) |
| Intermediate-high-risk (5 to <20%) | 325 (49.8) |
| High (≥20%) | 15 (2.3) |
| Statin eligibility* | 15 (2.3) |

Legend: \* statin eligibility criteria by each model is detailed in the Supplement. 95% CI calculated using one sample proportions test.

**Figure 1:**
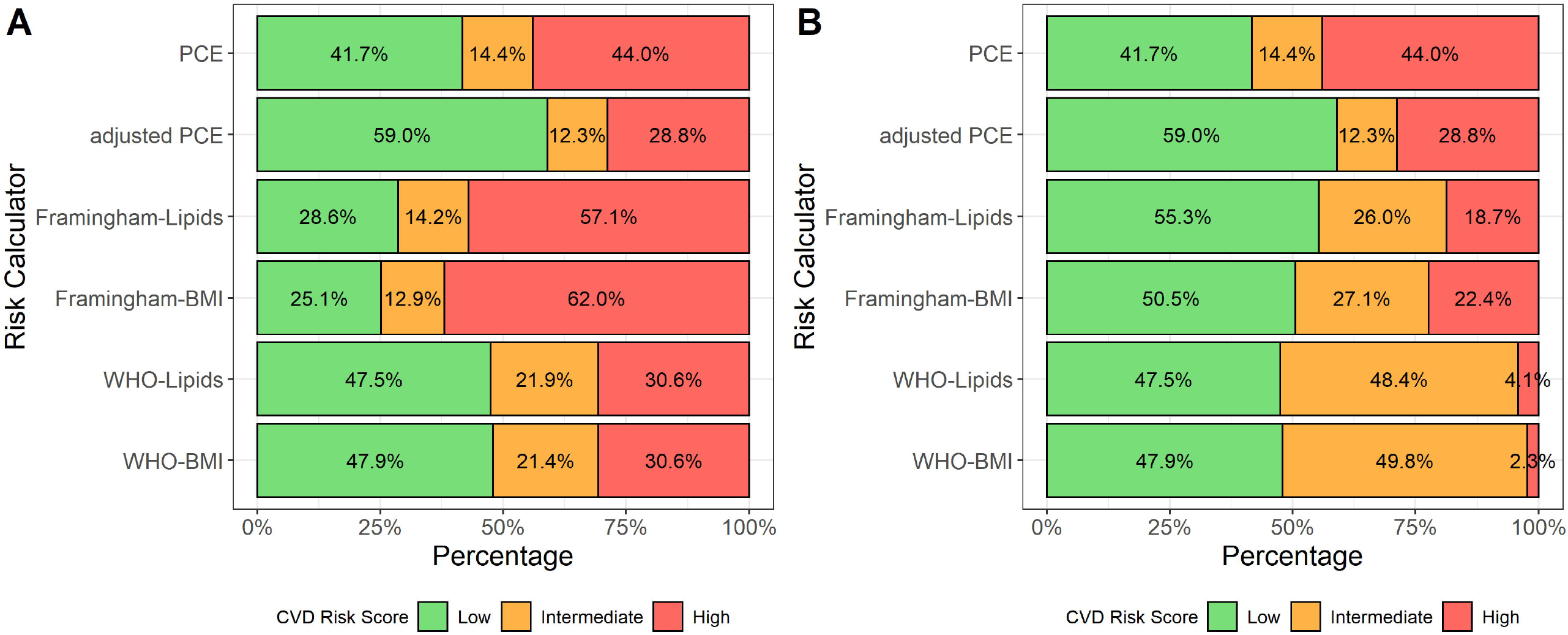
Predicted 10-year CVD risk categorizations by model. Legend: Figure shows proportion of cohort categorized as low, intermediate, or high-risk. Panel A uses a uniform threholds for low, intermediate, and high-risk: <5%, 5 to 7.5%, and ≥ 7.5%. Panel B uses model specific thresholds for low, intermediate, and high-risk: PCE <5%, 5 to 7.5%, ≥ 7.5%; adjusted PCE <5%, 5 to 7.5%, ≥ 7.5%; Framingham-Lipids <10%, 10 to 20%, ≥ 20%; Framingham-BMI <10%, 10 to 20%, ≥ 20%; WHO-Lipids <5%, 5 to 20%, ≥ 20%; WHO-BMI <5%, 5 to 20%, ≥ 20%.

Using model specific thresholds resulted in similarly wide variability in risk categorization (Table 2, Fig 1B). The percent of the cohort categorized as high-risk ranged from 2.3% in WHO-BMI to 44.0% in PCE (χ^2^ = 712, p value < 0.001). Under model specific thresholds, 70 participants had discordant scores, with the most common pattern as high risk by PCE and low risk by another model (68 out of 70 discordant participants).

### Risk Factor Distribution in High-Risk Category and Statin Eligibility

The risk factor distribution of age, sex, comorbidities, SBP, total cholesterol, and HDL cholesterol for participants with high 10-year CVD risk are summarized in Table 3. The median age ranged from 60 to 66, and percent female was 54.3% to 64.1%, reflecting the overall cohort sex breakdown. Diabetes and current smoking were not common (< 20% and < 15%, respectively) in the high-risk groups. However, SBP was relatively high. Treated SBP, or participants taking antihypertensive medications, ranged from a median of 152 to 162 mmHg, and untreated SBP ranged from a median of 142 to 158 mmHg. Total cholesterol was also high, ranging from a median of 197 to 202 mg/dL.

**Table 3:**
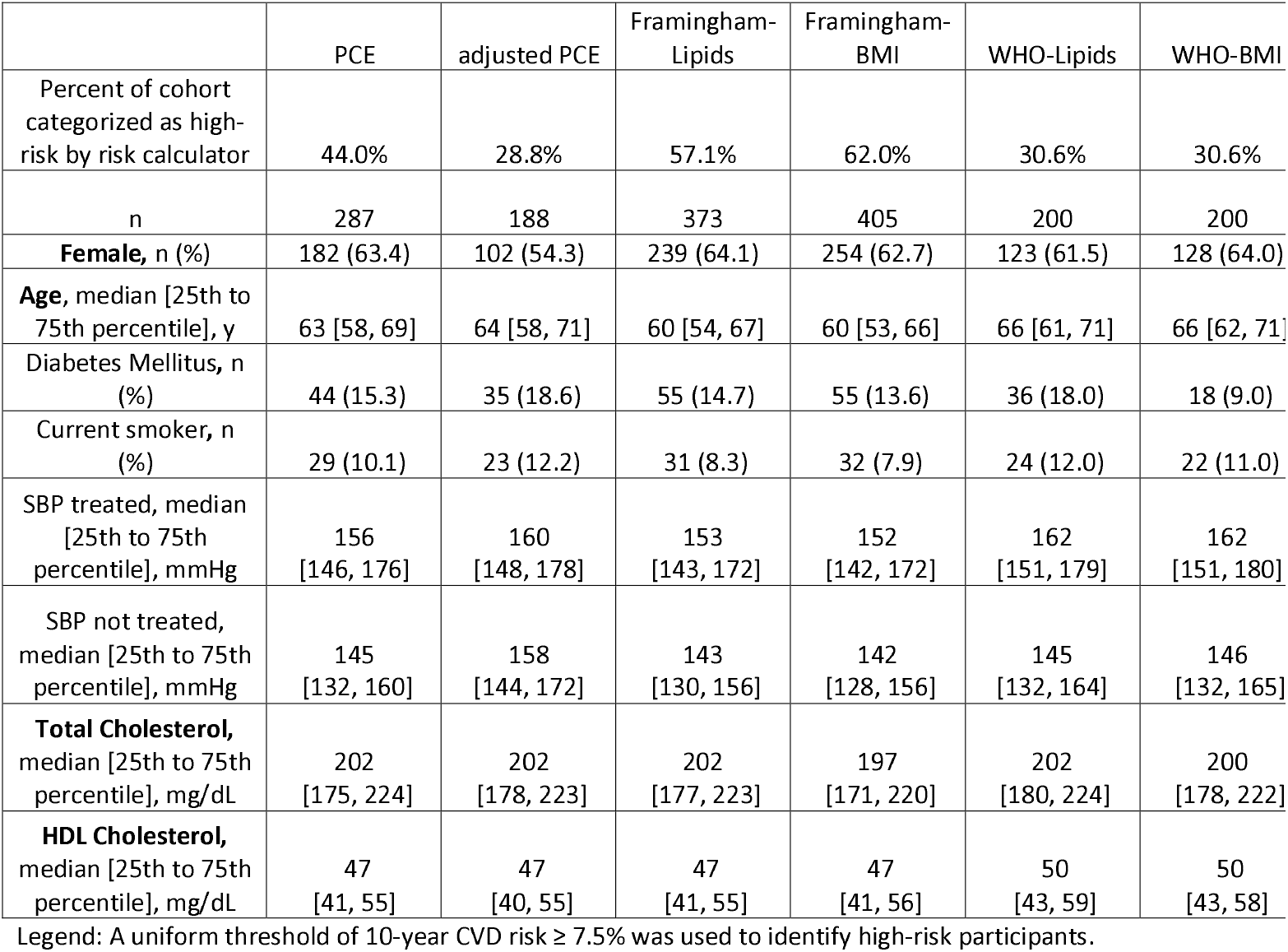
Risk factor distribution in high-risk category, using uniform thresholds.

|  | PCE | adjusted PCE | Framingham-Lipids | Framingham-BMI | WHO-Lipids | WHO-BMI |
| --- | --- | --- | --- | --- | --- | --- |
| Percent of cohort categorized as high-risk by risk calculator | 44.0% | 28.8% | 57.1% | 62.0% | 30.6% | 30.6% |
| n | 287 | 188 | 373 | 405 | 200 | 200 |
| **Female**, n (%) | 182 (63.4) | 102 (54.3) | 239 (64.1) | 254 (62.7) | 123 (61.5) | 128 (64.0) |
| **Age**, median [25th to 75th percentile], y | 63 [58, 69] | 64 [58, 71] | 60 [54, 67] | 60 [53, 66] | 66 [61, 71] | 66 [62, 71] |
| Diabetes Mellitus, n (%) | 44 (15.3) | 35 (18.6) | 55 (14.7) | 55 (13.6) | 36 (18.0) | 18 (9.0) |
| Current smoker, n (%) | 29 (10.1) | 23 (12.2) | 31 (8.3) | 32 (7.9) | 24 (12.0) | 22 (11.0) |
| SBP treated, median [25th to 75th percentile], mmHg | 156 [146, 176] | 160 [148, 178] | 153 [143, 172] | 152 [142, 172] | 162 [151, 179] | 162 [151, 180] |
| SBP not treated, median [25th to 75th percentile], mmHg | 145 [132, 160] | 158 [144, 172] | 143 [130, 156] | 142 [128, 156] | 145 [132, 164] | 146 [132, 165] |
| **Total Cholesterol**, median [25th to 75th percentile], mg/dL | 202 [175, 224] | 202 [178, 223] | 202 [177, 223] | 197 [171, 220] | 202 [180, 224] | 200 [178, 222] |
| **HDL Cholesterol**, median [25th to 75th percentile], mg/dL | 47 [41, 55] | 47 [40, 55] | 47 [41, 55] | 47 [41, 56] | 50 [43, 59] | 50 [43, 58] |

Legend: A uniform threshold of 10-year CVD risk $\geq 7.5\%$ was used to identify high-risk participants.

Using model specific thresholds, statin eligibility varied from 2.3% (95% CI 1.4% to 3.7%) with WHO-BMI to 44.9% (95% CI 41.1% to 48.7%) with PCE (χ^2^ = 513, p value < 0.001) (Table 2).

## Discussion

Correctly identifying high-risk patients allows for targeted interventions for primary prevention of CVD and treatment of underlying risk factors. In the Haiti CVD Cohort, we found substantial variation in the proportion identified as high-risk using existing models, ranging from 28.8% to 62.0% across models using uniform thresholds. Anywhere from 2.3% to 44.9% of participants were eligible for statins, leading to very different treatment recommendations and public health implications depending on which prediction model is chosen.

In LMIC, comparison of predicted risk in global cohorts using multiple models shows similar heterogeneity to our findings. In Brazil, an analysis comparing PCE to Framingham-Lipids in an all male cohort revealed Framingham categorized 5.5% of men as high-risk, while PCE categorized none as high-risk (14). In a cohort of HIV-infected patients in Botswana, PCE classified 14.1% as high risk while Framingham classified only 2.6% (15).

Existing CVD risk models are built on largely Caucasian populations, and may not be accurate for a majority black LMIC population like Haiti or many sub-Saharan African countries. Traditional methods using Cox proportional hazards may overfit the data on small subgroups like African Americans, leading to inaccurate predictions, and require assumptions about proportional hazards which may not be true (13). Newer statistical techniques, like machine learning, may avoid these limitations and integrate a larger breadth of data (16). Cohorts representative of LMIC with hard CVD outcomes are also needed to supply accurate underlying data.

There is an urgent need to design and validate CVD risk prediction tools in LMICs that include locally relevant risk factors reflecting disease pathology, and usability in low-resource settings. While ischemic heart disease accounts for the majority of CVD in high income countries (HIC), nonatherosclerotic stroke, hypertensive heart disease, and nonischemic cardiomyopathies are more common in LMIC (17). In our cohort, examining the high-risk group across models showed hypertension was relatively common, while diabetes and smoking were not. Designing new CVD risk models will also require a focus on usability. Lipids are not routinely available in many places (18), making non-lab based methods such as those using BMI more feasible. While online CVD risk calculators are widely available in HIC, lack of reliable internet and friction of integration into busy workflows suggest paper based wallcharts, such as produced by the WHO, may work better in LMIC.

To achieve desired health outcomes, CVD risk prediction must be translated into successful action, involving multisector action from health systems, health care providers, and patients. Statin accessibility is low in many LMICs. Based on the WHO Health Action International survey, statins are not on the essential medicine list of 34% of countries, including Haiti (19,20). In Haiti, a 2011 survey showed atorvastatin and simvastatin were available in retail pharmacies, but rarely in public or nonprofit pharmacies, and expensive (21). The lowest paid government worker would need 2.6 days wages to pay for a 1 month supply of statins if bought from a public sector pharmacy, and 13.7 day wages if brought from a retail pharmacy (21). Lower availability and affordability of essential CVD meds have been associated with higher risk of major adverse cardiovascular events and mortality (HR 1.25, 95% CI 1.08 to 1.50) (22).

Strengths of this study include it’s design as a population-based cohort, rigorous BP measurement, and standardized lipid measurement. Limitations include the exclusion of young participants < 40 for whom traditional CVD risk models do not apply, and the lack of hard CVD outcomes to conduct predicted versus observed comparisons.

In summary, across six commonly used CVD risk prediction models, there was substantial variation in identification of high-risk participants using both uniform, and model specific thresholds. Locally relevant CVD risk prediction models are needed in LMIC, combined with health systems strengthening to increase treatment availability and affordability.

## Supporting information

STROBE checklist

## Data Availability

Researchers who provide a methodologically sound proposal may have access to a subset of deidentified participant data, with specific variables based on the proposal. Proposals should be directed to the principal investigator at. To gain access, data requestors will need to sign a data access agreement. Data are available following publications through 3 years after publication and will be provided directly from the PI.

## List of abbreviations

aPCE: adjusted pooled cohort equations
BMI: body mass index
BP: blood pressure
CVD: cardiovascular disease
DBP: diastolic blood pressure
GHESKIO: Groupe Haïtien d’Etude du Sarcome de Kaposi et des Infections Opportunistes clinics
IQR: interquartile range
LMIC: low middle income countries
PCE: pooled cohort equations
SBP: systolic blood pressure
SCORE: systematic coronary risk evaluation model
WHO: World Health Organization

## Declarations

### Ethics approval and consent to participate

This study was approved by institutional review boards at Weill Cornell Medicine and Groupe Haitien d’Etude du Sarcome de Kaposi et des Infections Opportunistes (GHESKIO) (record number 1803019037), with written participant consent.

### Consent for publication

Not applicable

### Author Contributions

This study was conceived by LDY and MLM. Project administration, and data curation were completed by LDY, JLP, VR, JWP, MLM. Formal analysis was completed by LDY. Investigation, methodology, and interpretation were completed by LDY, JLP, VR, MT, JWP, MMS, and MLM. LDY and MLM wrote the initial draft. All authors participated in reviewing and editing the manuscript.

All authors have read, and confirm that they meet, ICMJE criteria for authorship.

## Acknowledgements

We acknowledge the valuable input from the Haitian College of Cardiology, the community health workers who help with data collection, the data management team (Stephano St-Preux, Olga Tymejczyk, Miranda Metz), and the study participants for entrusting us with their care.

## Author access to data

LDY and MLM had full access to all the data in the study and take responsibility for the integrity of the data and the accuracy of the data analysis. LDY conducted the data analysis.

