## Supplemental Materials for "Cardiovascular Disease Risk Prediction Models in Haiti: Implications for Primary Prevention in Low-Middle Income Countries"

### Supplemental Figure 1

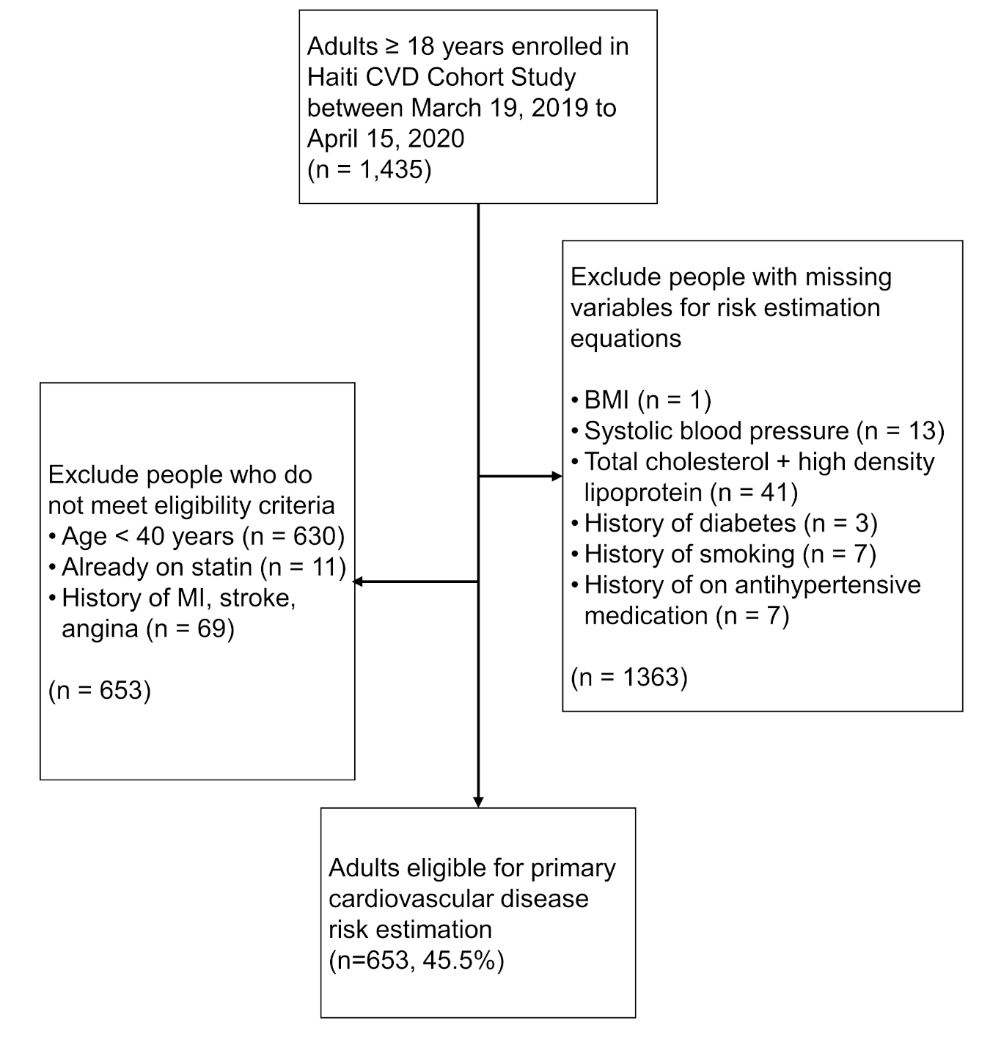

### Supplemental Table 1

| **Condition** | **Clinical or laboratory measurement** |
| --- | --- |
| Hypertension | - Study physician diagnosed patient with hypertension based on history and physical - Patient reports taking medication for high blood pressure - Average systolic blood pressure of last two out of three measurements ≥ 140 mmHg OR average diastolic blood pressure of last two out of three measurements ≥ 90 mmHg |
| Hypercholesterolemia | - Study physician diagnosed patient with hypercholesterolemia based on history and physical - Patient reports taking medication for hypercholesterolemia - non-HDLc 190-219 mg/L, OR LDLc ≥160 mg/dL ^1^. To convert LDLc to millimoles per liter, multiply by 0.0259 |
| Diabetes | - Study physician diagnosed patient with diabetes based on history and physical - Patient reports taking medication for diabetes - random glucose level ≥ 200 mg/dL OR fasting glucose level ≥ 126 mg/dL |

### Methodology for CVD risk prediction application

CVD risk prediction models are largely based on Cox proportional hazard models, developed using data from large population cohorts. The exception to this is the adjusted Pooled Cohort Equation (PCE) model developed by Yadlowsky et al ^2^, which uses newer statistical methods (elastic net regularization selection of logistic equations) to avoid the specific weaknesses of the original PCE derivation process: overfitting in small subpopulations like black adults, and violation of the proportional hazards assumption which affects the accuracy of risk estimates among subgroups.

The general process of calculating an individual’s 10 year CVD risk is as follows:

Step 1: Find underlying equation and coefficients from published literature ^1–5^

Step 2: Construct equations in R, building off previously published work ^2^

Step 3: Calculate CVD risk for each individual in cohort

Step 4: As a quality check, take a random 10% sample and manually calculate CVD risk using publicly available online calculators for PCE, adjusted PCE, Framingham Lipids, and Framingham BMI. Used publicly available wallcharts for WHO Lipids and WHO BMI. Compared manually calculated scores to values obtained in Step 3, and correct errors in equations as needed

The specific coefficients used from the published literature are detailed in Supplemental Tables 2-4. For the WHO Lipids and WHO BMI risk calculation, while the coefficients and equations are publicly available, Kaptoge et al ^5^ used Global Burden of Disease incident statistics to derive two rescaling factors for each world region, and then present a recalibrated 10 year CVD risk. However, these rescaling factors are not available publicly. Instead, we used the published WHO wallcharts to write a program in R (modeled off the whoishRisk package) that takes the inputs of sex, diabetes, smoking status, age, systolic blood pressure, and either lipids or BMI, and outputs the recalibrated risk based on the wallchart.^5^

#### Supplemental Table 2: Pooled Cohort Equation coefficients

|  | **Coefficients** | | | |
| --- | --- | --- | --- | --- |
|  | Men | | Women | |
| **Independent variables** | White | African American | White | African American |
| Ln Age | 12.344 | 2.469 | –29.799 | 17.114 |
| Ln Age, squared | NA | NA | 4.884 | N/A |
| Ln Total cholesterol (mg/dL) | 11.853 | 0.302 | 13.54 | 0.94 |
| Ln Age × Ln Total Cholesterol | –2.664 | N/A | –3.114 | N/A |
| Ln HDL-C (mg/dL) | –7.990 | –0.307 | –13.578 | –18.920 |
| Ln Age × Ln HDL-C | 1.769 | N/A | 3.149 | 4.475 |
| Ln Treated Systolic BP (mm Hg) | 1.797 | 1.916 | 2.019 | 29.291 |
| Ln Age × Ln Treated Systolic BP | NA | NA | N/A | –6.432 |
| Ln Untreated Systolic BP (mm Hg) | 1.764 | 1.809 | 1.957 | 27.82 |
| Ln Age × Ln Untreated Systolic BP | NA | NA | N/A | –6.087 |
| Current Smoker (1=Yes, 0=No) | 7.837 | 0.549 | 7.574 | 0.691 |
| Ln Age × Current Smoker | –1.795 | N/A | –1.665 | N/A |
| Diabetes (1=Yes, 0=No) | 0.658 | 0.645 | 0.661 | 0.874 |
| Mean (Coefficient × Value) | 61.18 | 19.54 | –29.18 | 86.61 |
| S_10_, or baseline survival estimate at 10 years | 0.9144 | 0.8954 | 0.9665 | 0.9533 |

Legend: 10 year risk of CVD event= $1-{S_{10}}^{exp(\sum\beta_{x}-mean)}$, from Goff et al, 2013 ^3^.

#### Supplemental Table 3: Adjusted Pooled Cohort Equation coefficients

|  | **Coefficients** | |
| --- | --- | --- |
| **Independent variables** | Men | Women |
| Age | 0.0642 | 0.106501 |
| Black race (1/0 for black/white) | 0.482835 | 0.43244 |
| Systolic blood pressure (mm Hg) squared | −0.000061 | 0.000056 |
| Systolic blood pressure | 0.03895 | 0.017666 |
| Taking blood pressure medication (1/0 for yes/no) | 2.055533 | 0.731678 |
| Diabetes mellitus (1/0 for yes/no) | 0.842209 | 0.94397 |
| Current smoker (1/0 for yes/no) | 0.895589 | 1.00979 |
| Ratio of total cholesterol (mg/dL) to high-density lipoprotein cholesterol (mg/dL) | 0.193307 | 0.151318 |
| Age if black (0 if not) | NA | −0.008580 |
| Systolic blood pressure if taking blood pressure medication (0 if not) | −0.014207 | −0.003647 |
| Systolic blood pressure if black (0 if not) | 0.011609 | 0.006208 |
| Black race and taking blood pressure medication (1/0 for yes/no) | −0.119460 | 0.152968 |
| Age × systolic blood pressure | 0.000025 | −0.000153 |
| Black race and diabetes mellitus (1/0 for yes/no) | −0.077214 | 0.115232 |
| Black race and current smoker (1/0 for yes/no) | −0.226771 | –0.092231 |
| Ratio of total cholesterol to high-density lipoprotein cholesterol if black | −0.117749 | 0.070498 |
| Systolic blood pressure if black and taking blood pressure medication (0 if not) | 0.00419 | −0.000173 |
| Age × systolic blood pressure if black (0 if not) | −0.000199 | −0.000094 |
| Mean (intercept) | −11.679980 | −12.823110 |

Legend: 10 year risk of CVD event=$\frac{1}{1+exp\left( -sum of terms \right)}$, from Yadlowsky et al, 2018 ^2^.

#### Supplemental Table 4: Framingham Lipids and Framingham BMI coefficients

|  | **Coefficients** | |
| --- | --- | --- |
| **Independent variables** | Men | Women |
| **Framingham Lipids** |  |  |
| Log of Age | 3.06117 | 2.32888 |
| Log of Total Cholesterol | 1.1237 | 1.20904 |
| Log of HDL Cholesterol | -0.93263 | -0.70833 |
| Log of SBP if not treated | 1.93303 | 2.76157 |
| Log of SBP if treated | 1.99881 | 2.82263 |
| Smoking | 0.65451 | 0.52873 |
| Diabetes | 0.57367 | 0.69154 |
| Mean (Coefficient × Value) | 23.9802 | 26.1931 |
| S_10_, or baseline survival estimate at 10 years | 0.88936 | 0.95012 |
| **Framingham BMI** |  |  |
| Log of Age | 3.11296 | 2.72107 |
| Log of Body Mass Index | 0.79277 | 0.51125 |
| Log of SBP if not treated | 1.85508 | 2.81291 |
| Log of SBP if treated | 1.92672 | 2.88267 |
| Smoking | 0.70953 | 0.61868 |
| Diabetes | 0.5316 | 0.77763 |
| Mean (Coefficient × Value) | 23.9388 | 26.0145 |
| S_10_, or baseline survival estimate at 10 years | 0.88431 | 0.94833 |

Legend: 10 year risk of CVD event = $1-{S_{10}}^{exp(\sum\beta_{x}-mean)}$, from D’Agostino et al, 2008 ^4^.

### Supplemental Table 5: Statin eligibility criteria

Each CVD risk prediction model has separate criteria for when statins are recommended for primary prevention of CVD. In general, statins are recommended for high risk groups.

| Risk prediction model | Statin eligibility for primary prevention |
| --- | --- |
| PCE, adjusted PCE^1^ | - LDL cholesterol ≥ 190 mg/dL - Diabetes and LDL cholesterol ≥ 70 mg/dL - Calculated 10-year cardiovascular disease risk of 7.5% or greater, based on the pooled cohort equations and LDL cholesterol ≥ 70 mg/dL |
| Framingham Lipids, Framingham BMI ^6^ | - LDLc ≥ 190 mg/dL - Diabetes (self-reported or fasting glucose ≥ 126 mg/dL) and LDL cholesterol ≥ 100 mg/dL - Combination of calculated 10-year cardiovascular disease risk (using the Framingham risk calculator) and LDL cholesterol levels:   - Risk of 20% or greater and LDL cholesterol ≥ 100 mg/dL   - Risk of 10 to 20% and LDL cholesterol ≥ 130 mg/dL with 2 or more risk factors [smoking, hypertension, high-density lipoprotein (HDL) cholesterol < 40 mg/dL, myocardial infarction or angina in first degree relative before age 50, and age (45 years or older for men, 55 years or older for women)]   - Risk < 10% and LDL cholesterol ≥ 160 mg/dL with 2 or more risk factors |
| WHO Lipids, WHO BMI ^7^ | - Calculated 10-year cardiovascular disease risk ≥ 20% |
